# Pooling RT-PCR test of SARS-CoV-2 for large cohort of “healthy” and infection-suspected patients: A prospective and consecutive study on 1,000 individuals

**DOI:** 10.1101/2020.05.04.20088146

**Authors:** Yosuke Hirotsu, Makoto Maejima, Masahiro Shibusawa, Yuki Nagakubo, Kazuhiro Hosaka, Kenji Amemiya, Hitomi Sueki, Miyoko Hayakawa, Hitoshi Mochizuki, Masao Omata

## Abstract

**Background:** SARS-CoV-2 testing reagents are expected to become in short supply worldwide. However, little is unknown whether the pooling strategy detects SARS-CoV-2 with accuracy.

**Method:** To validate the feasibility of pooling samples, serial dilution analysis and spike-in experiment were conducted using synthetic DNA and nucleic acids extracted from SARS-CoV-2 positive and negative patients. Furthermore, we studied a total of 1,000 individuals, who were 667 “healthy” (195 healthcare workers and 472 hospitalized patients with other disorders than COVID-19 infection) individuals and 333 infection-suspected patients with cough and fever, were tested.

**Results:** Serial dilution analysis showed the limit of detection of around 10-100 copies according to National Institute of Infectious Diseases, Japan. Spike-in experiment demonstrated RT-qPCR detect positive signal in pooling samples of SARS-CoV-2 negative and positive patient at the 5-, 10-, 20-fold dilution. By screening with pooling strategy by the end of April, 2020, there are 12 COVID-19 patients in 333 infection suspected patients (3.6%) and zero in 667 “healthy”. We obtained these results with total running 538 times (instead of 1,000 times) by pooling strategy.

**Conclusion:** Pooling samples is feasible for saving test reagents and detecting SARS-CoV-2 in clinical setting to prevent the spread of the virus and nosocomial transmission.

## Introduction

A new emergent coronavirus, severe acute respiratory syndrome coronavirus 2 (SARS-CoV-2), outbreak in late 2019 in Wuhan, China, spread across the world in a several months [1, 2]. SARS-CoV-2 has caused high number of infected patients with coronavirus disease 2019 (COVID-19) and death at 2.3% in China and 7.2% in Italy [3]. SARS-CoV-2 have been spread by infected people who have mild or no symptoms [4]. The World Health Organization (WHO) has sounded a warning to the world, declared it a pandemic, and announced the need for a testing system for suspected patients who infected with SARS-CoV-2. The development of a testing system for SARS-CoV-2 is an urgent issue.

The explosive increase in COVID-19 cases has depleted medical resources to protect against infection [5]. For instance, face shields, infection control gowns, N95 masks, surgical masks, and personal protective equipment (PPE) are in short supply at hospital [6]. As a result, the risk of infection among medical staffs and nosocomial infection is expected to be high. To prevent the spread the virus, the healthcare workers carefully clean the environment around the hospital room [7]. In addition, screening tests for healthcare workers, ambulatory patients, hospitalized patients and asymptomatic cases are important for early detection of SARS-CoV-2 and prevention of nosocomial infections. Prophylactic testing can help protect healthcare workers and patients in clinical setting.

As COVID-19 has spread, tens of thousands of tests have been performed worldwide in a day. Real-time reverse transcription-PCR (RT-PCR) is now widely used for the detection of SARS-CoV-2 [8]. RT-PCR was conducted as following steps: viral RNA extraction, reverse transcription reaction, and real-time PCR detection with primers and fluorescent probe. It is expected that the supply of reagents will be depleted as growing global demands for testing SARS-CoV-2. In particular, virus nucleic acid extraction reagents and enzyme-containing reagents for real-time PCR are necessary to perform the test. In the U.S., testing has been stopped in some areas due to lack of testing [9]. It is important to consider how to conduct the RT-PCR testing with accuracy while saving reagents

Analysis of pooled samples is time- and cost-saving method for diagnosis of infectious diseases. Robert *et al*. successfully identified syphilis from pooled samples at 1943 [10]. Pooling strategies have also been used for the detection of other pathogens including hepatitis B virus, hepatitis C virus, human immunodeficiency virus, Chlamydia trachomatis and Neisseria gonorrhoeae [11–15]. Pooling strategy was conducted for screening SARS-CoV-2 in U.S. [9, 16]. In this study, we validated the pooling strategy for detecting SARS-CoV-2 using multiple nasopharyngeal swabs. This method would help us to save time and reagents under short supply conditions and prevent delays in reporting results. This could provide the prevalence of ongoing infection in 667 healthy in our distinct, west of Tokyo.

## Materials and Methods

### Samples of patients and medical staffs

We collected nasopharyngeal swabs between March 11 and April 28, 2020 at Yamanashi Central Hospital. All samples were obtained with cotton swab and universal transport media (Copan, Murrieta, CA). To screen whether medical staffs and hospitalized patients were infected with SARS-CoV-2, we tested a total of 1,000 samples from 1,000 individuals. By pooling strategy, we tested a total of 538 samples (445 individuals and 93 pools). One pooled batch was made of 5 to 10 samples. This study was approved by the Institutional Review Board at Yamanashi Central Hospital and complied with Declaration of Helsinki principles.

### Viral nucleic acid extraction

Total nucleic acid was automatically isolated from nasopharyngeal swabs using the MagMax Viral/Pathogen Nucleic Acid Isolation Kit (ThermoFisher Scientific, Waltham, MA) on automated machine KingFisher Duo Prime according to the manufacturer’s protocol. Briefly, we added 400 μL of viral transport media, 10 μL of Proteinase K, 530 μL Binding Solution, 20 μL Total Nucleic Acid Binding Beads, 1mL Wash Buffer, and 1mL or 0.5mL of 80% Ethanol to each well of a Deep-well 96-well plate. 100 μL of Elution solution was added to Elution Strip. Total nucleic acids were stored at −80 °C until further RT-PCR analysis.

### One-step real-time quantitative RT-PCR (RT-qPCR)

The protocol were designed by National Institute of Infectious Diseases (NIID), Japan [17]. To detect SARS-CoV-2, we performed one-step real-time quantitative RT-PCR according to the NIID protocol with minor modification (version 2.7) [17]. The primer/probe set testes two sites (N1 and N2) of the *N* gene of SARS-CoV-2 [17, 18].

For N1 detection with the NIID assay, the reaction mixture comprised 5 μL of 4× TaqMan Fast Virus 1-Step Master Mix, 1.2 μL of 10 μM forward primer, 1.6 μL of 10 μM reverse primer, 0.8 μL of 5 μM probe, 6.4 μL of nuclease-free water (Thermo Fisher Scientific), and 5 μL of sample in a 20 μL total volume. For N2 detection, the reaction mixture comprised 5 μL of 4× TaqMan Fast Virus 1-Step Master Mix, 1.0 μL of 10 μM forward primer, 1.4 μL of 10 μM reverse primer, 0.8 μL of 5 μM probe, 6.8 μL of nuclease-free water, and 5 μL of sample in a 20 μL total volume. For the internal positive control, the human ribonuclease P 30 subunit (*RPP30*) gene was used [18]. The RT-PCR assays were conducted on a StepOnePlus Real-Time PCR Systems (Thermo Fisher Scientific) with the following cycling conditions: 50°C for 5 min for reverse transcription, 95°C for 20 s, and 45 cycles of 95°C for 3 s and 60°C for 30 s. The threshold line was set at 0.2. The threshold cycle (Ct) value was assigned to each PCR reaction and the amplification curve was visually assessed. The absolute copy number of viral loads was determined using serial diluted DNA control targeting *N* gene of SARS-CoV-2 (Integrated DNA Technologies, Coralville, IA).

### Serial dilution assay using SARS-CoV-2 DNA plasmid control

We purchased the SARS-CoV-2 DNA plasmid control (Integrated DNA Technologies, catalog #10006625), which consists of 200,000 copies/μL containing the complete *N* gene [18]. We prepared a serial dilution of the plasmid control (100,000, 10,000, 1,000 and 100 copies) using nuclease-free water (Thermo Fisher Scientific) to assess the limit of detection. Serial diluted plasmid was mixed with total nucleic acid extracted from SARS-CoV-2-negative healthy individuals at a ratio of 1:9. Plasmid control with a 10-fold dilution of the final concentration were analyzed by RT-PCR.

### Spike-in assay using COVID-19 and non-COVID-19 patients

Representative three nucleic acids extracted from COVID-19 patients were used for spike-in analysis. These samples contained high, intermediate and low virus loads. Using these three samples, we examined how far SARS-CoV-2 could be detected when multiple samples were pooled. SARS-CoV-2 positive and negative samples were mixed in ratios of 1:4, 1:9, and 1:19. As a result, pooled samples of 5-, 10- and 20-fold dilution were created, respectively.

## Results

### Serial dilution assay using plasmid control and SARS-CoV-2 negative

RT-PCR for SARS-CoV-2 was conducted based on protocols developed by the NIID in Japan [17]. This assay targets two sites of the *N* gene of SARS-CoV-2. We previously conducted serial dilution experiment with positive control plasmid and observed primer/probe targeting N2 site has more sensitive than that of N1 site [18]. To assess the feasibility of pooling samples, we diluted plasmid control containing the *N* gene of SARS-CoV-2 with total nucleic acids extracted from SARS-CoV-2-negative healthy individuals. First, we diluted synthetic plasmid control with different concentrations (100,000, 10,000, 1,000 and 100 copies). These different diluted samples were mixed with nucleic acids extracted from nasopharyngeal swabs of SARS-CoV-2-negative healthy individuals at the ratio of 1:9 ratio. As a result, the expected amount of inputs for 10,000, 1,000, 100 and 10 copies were analyzed by RT-PCR (Figure 1A). The results showed primer/probe targeting N1 site detected 10,000 and 1,000 copies of plasmid, but did not detected 100 and 10 copies; whereas N2 site detected down to 100 copies (Figure 1B). These results suggested the limit of detections were 100-1000 copies and 10-100 copies by N1 and N2 site, respectively.

**Figure 1.**
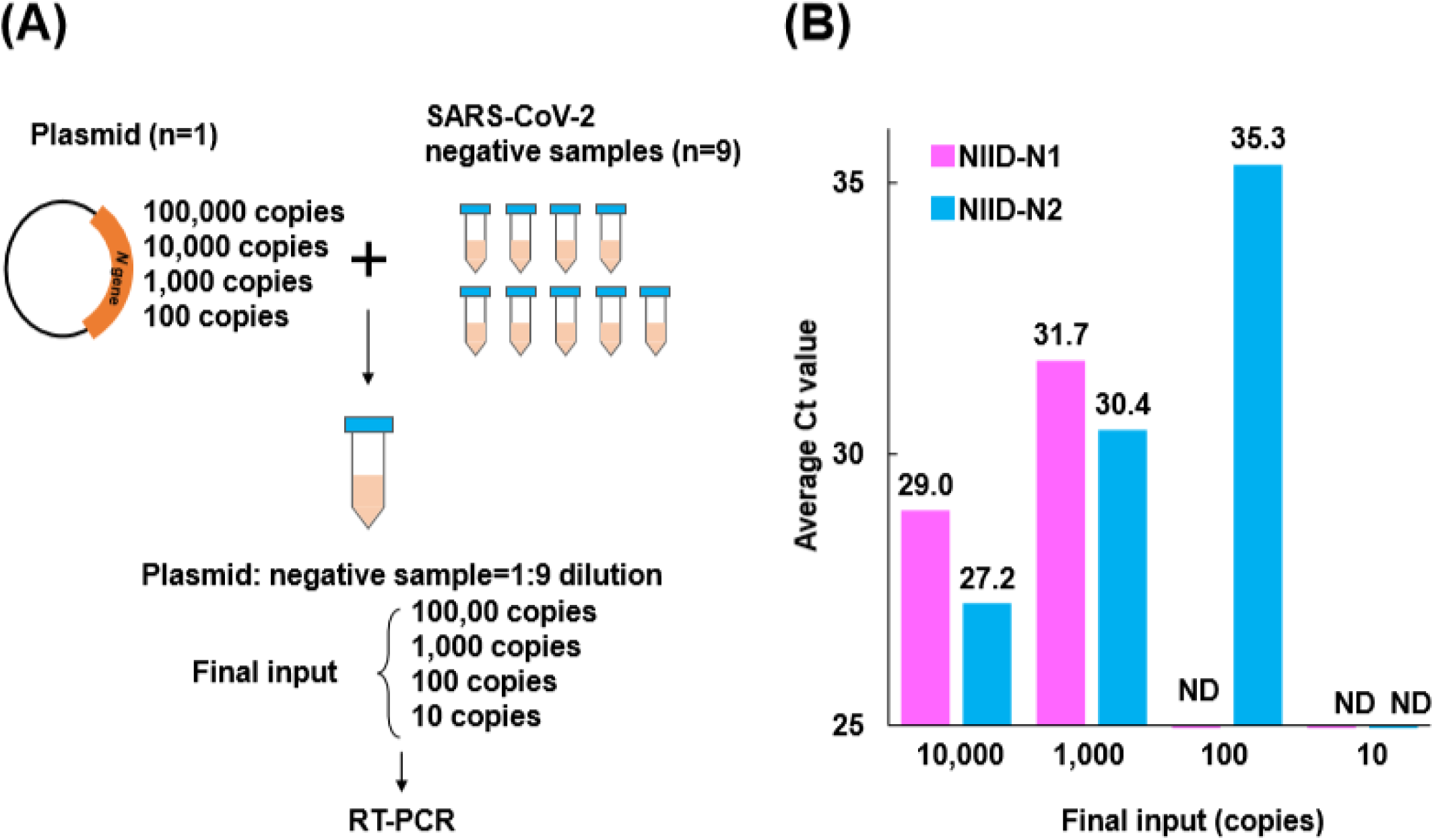
Serial dilution analysis with synthetic plasmids and nasopharyngeal swabs from non-COVID19 individuals. **(A)** Scheme of preparing serial dilution solutions. The synthetic plasmid containing *N* gene of SARS-CoV-2 were diluted at the concentration of 100,000, 10,000, 1,000 and 100 copies. These each plasmid solution were 10-fold diluted with nucleic acids extracted from nasopharyngeal swabs from non-COVID19 patients (n=9). RT-PCR analysis have already validated there are no amplification was observed in non-COVID-19 patients in advance. RT-PCR analysis were conducted using serial dilution solution with final input copies numbers (ranged 10 to 10,000). **(B)** Average threshold cycle (Ct) were determined by RT-PCR. Two sets of primers and probes (pink, NIID-N1; blue, NIID-N2) were used according to NIID, Japan. The experiment was conducted three times in duplicate.

### Spike-in assay using SARS-CoV-2 positive and negative samples

We pooled samples from the SARS-CoV-2 positive patient and negative health individuals. Samples with three different concentrations of SARS-CoV-2 viral loads were diluted in negative samples at a ratio of 1:4, 1:9, and 1:19 (Figure 2A). As a result, samples were prepared at 5-, 10- and 20-fold dilution, respectively. These samples were subjected to RT-qPCR. RT-qPCR-showed the original viral load in the undiluted sample was calculated to be 7.8 log_10_ (high viral load), 5.8 log_10_ (intermediate), and 1.3 log_10_ (low) at the N1 site (Figure 2B), and 9.3 log_10_ (high), 6.8 log_10_ (intermediate), and 3.6 log_10_ (low) at the N2 site (Figure 2C). The primer/probe targeting N1 site was detectable with high and moderate viral load samples, but not with low viral load when diluted samples (Figure 2B). In contrast, N2 site assay detected using high, moderate and low viral load (Figure 2C). These results showed N2 site is feasible for analyzing the pooling samples rather than N1.

**Figure 2.**
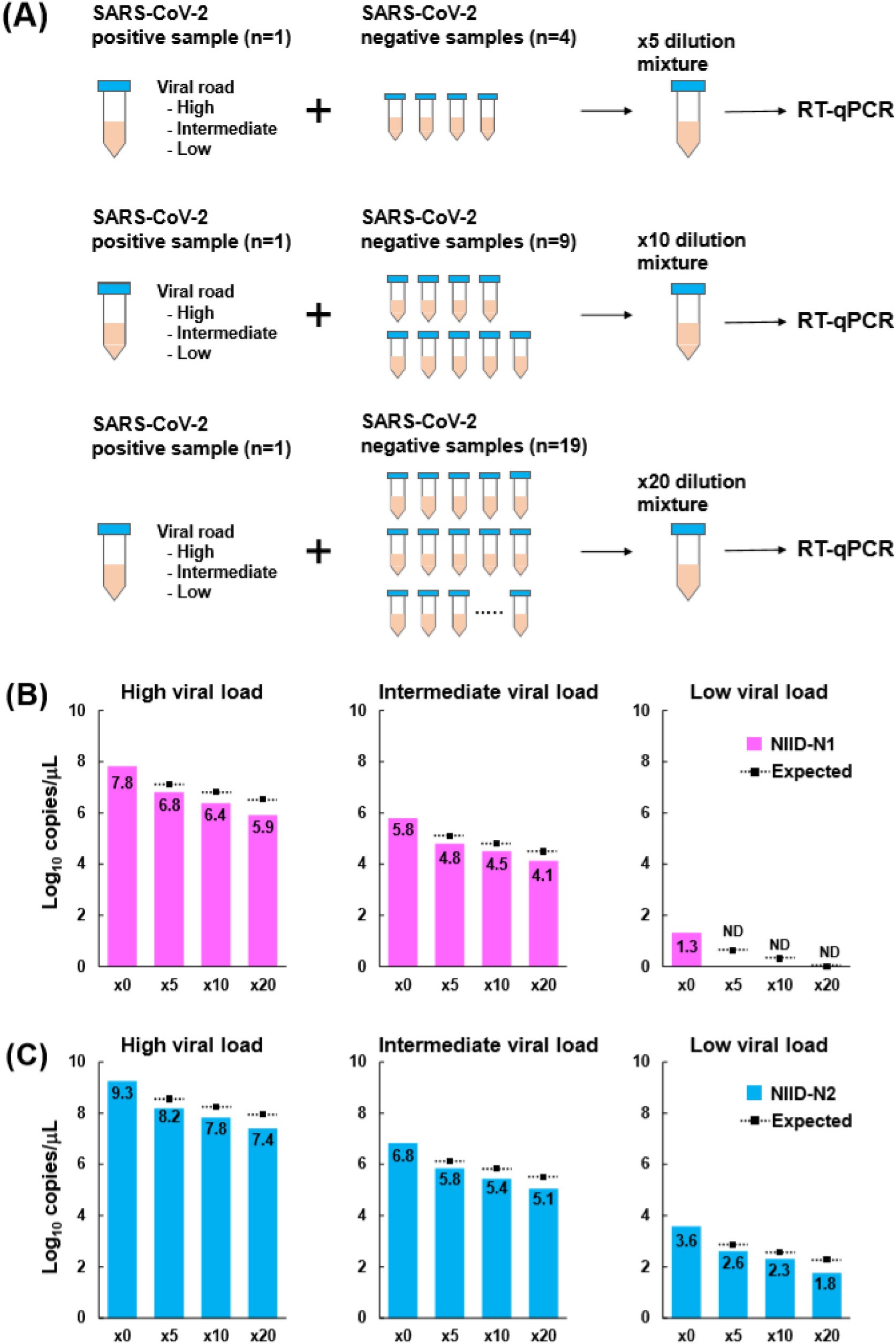
Spike in assay using SARS-CoV-2 positive and negative nasopharyngeal swabs from patients. **(A)** Scheme of preparing spike in solutions. SARS-CoV-2 positive samples have high, intermediate and low viral copies. These different viral load were diluted with 4, 9 and 19 SARS-CoV-2 negative samples. The final solution was made at the ×5, ×10 and ×20 dilution and used for RT-qPCR analysis. **(B-C)** RT-qPCR analysis determined the copy numbers in spike in solution. The assay was used with NIID-N1 (B) and NIID-N2 (C).Bar plot shows the copy number (log_10_ copies/μL) in original (×0) and diluted (×5, ×10 and ×20) samples. Dot line shows the copy numbers theoretically reduced by dilution.

Based on the concentration of the undiluted sample, we compared the data between the number of copies expected by dilution and the observed number of copies. The observed copy number was estimated lower than the expected virus load in N1 and N2 sites (Figure 2B and 2C). The decline in the number of copies according to dilution was close to the expected value, indicating there were low effects of reaction inhibitors and contaminant in the swabs on the measurements using pooling samples.

### SARS-CoV-2 test for COVID-19 suspected patients, healthcare workers and hospitalized patients

From March 11 to April 28, 2020, we studies a total of 1,000 individuals including 333 COVID-19 infection suspected patients, 195 healthcare workers, 472 hospitalized patients with disorder other than COVID-19 (Table 1). RT-qPCR showed the prevalence of COVID-19 was 3.6% (12/333) of infection-suspected patients and none in both healthcare workers and hospitalized patients in our distinct (Table 1).

**Table 1.** Prevalence of COVID-19 in infection-suspected patients and screening group

|  | Suspected | Screening Group |  | Total |
| --- | --- | --- | --- | --- |
|  | COVID-19 Suspected Patients* | Healthcare Workers | Patients Hospitalized other than COVID-19 |  |
| Number of patients | 333 | 195 | 472 | 1,000 |
| COVID-19 positive/ negative (%) | 12 (3.6%)/ 321 (97.7%) | 0 (0%) / 195 (100%) | 0 (0%) / 472(100%) | 12 (1.2%) / 988 (98.8%) |
| Total tested samples | 333 | 91 | 114 | 538 |
| Individual | 333 | 65 | 47 | 445 |
| Polled | 0 | 26 | 67 | 93* |

\* This group includes the 12 COVID-19 confirmed patients.
\*\* 93 tested pooled samples from 555 individuals. A breakdown of the pooled samples: 66 (five samples in one batch), 3 (six samples), 4 (seven samples), 5 (eight samples), 11 (nine samples), 4 (ten samples).

We tested a total of 538 samples including 445 individuals and 93 pools (corresponding to 555 samples), which meant 46% of reagent was saved. To date, we prospectively followed up the COVID-19-negative hospitalized individuals and did not observe apparent symptom within the average 10-12 hospital stays, suggesting nosocomial infections could be prevented in our hospital.

## Discussion

Here, we shows the utility of polling strategy for detecting SARS-CoV-2. Serial dilution analysis with control plasmid DNA and nucleic acids extracted from healthy individuals showed the limit of detection was estimated as 10-100 using N2 site according to NIID protocol. The spike-in analysis with positive and negative SARS-CoV-2 nasopharyngeal swab specimen demonstrated that RT-qPCR detected SARS-CoV-2 even in pooled samples with viral loads. If we applied the pooling ten individual samples as one batch, we may obtain the results with only 100 times with a sensitivity loss of 1.3-1.5 log_10_ copies. These results show the utility of pooling strategy for screening healthcare workers and patients in hospital.

If the PCR efficiency is 100%, pooling of 5 samples theoretically decreases the viral loads from original value to 0.7 log_10_ (Ct value increases 2.3), 10 samples decreases to 1.0 log_10_ (Ct increases 3.3), and 20 samples decreases to 1.3 log_10_ (Ct increase 4.3). In case of N2 site of NIID assay, the limit of detection was estimated around 10-100 copies of virus loads from serial dilution assay using nucleic acids from non-COVID-19 healthy individuals. Therefore, our data suggests 5, 10 and 20 samples pooling strategy is feasible when the original viral load is 50-500, 100-1,000 and 200-2,000 copies, respectively. When we observed positive results in pooled samples, we have to retest the separately each in pooled sample. This strategy is effective to save the reagents and time for reporting SATR-CoV-2 testing. In U.S., 292 pools were screen from 2,888 individual samples and SARS-CoV-2 positive rate was 0.07% (2/2,888)

The viral load is peak at onset of symptom and then gradually decline in COVID-19 patients [19]. The median initial viral load reported as 6.17 log_10_ copies (range 4.18-7.13) in severe cases and 5.11 log_10_ copies (range 3.91-7.56) in mild cases [20]. These observed viral loads in COVID-19 will be detected in pooled samples. Hypothetically, asymptomatic COVID-19-positive individuals maybe have high viral loads before the 2-3 days of onset [20]. Screening the asymptomatic individuals would be have effect to prevent spreading the virus at hospital [21]. Collectively, pooling strategy may help to detect early and prevent the nosocomial infection in hospital. In addition, screening of inpatients, emergency admissions, and physicians and health care providers, as in this study, can be useful for infection control.

This pooling method would become powerful tool to detect asymptomatic “super spreaders” with heavy viral loads. In Japan, this super spreaders are key players for the increasing infection trend. Recently, a short report on apparently healthy individuals from university hospital in Tokyo reported 6% (4/67) prevalence on April 21, 2020 [22]. Fortunately, our distinct (100-150 km next to Tokyo) is not obviously invaded by this virus at the end of April, 2020. This method may help to elucidate the future trend of this infection and reveal the prevalence of COVID-19 patients without losing any accuracy in spite of fewer tested samples.

## Data Availability

Not applicable

## Acknowledgement.

We thank all of the medical and ancillary hospital staff and the patients for consenting to participate.**Financial support**. This study was supported by a Grant-in-Aid for the Genome Research Project from Yamanashi Prefecture (to M.O. and Y.H.), the Japan Society for the Promotion of Science (JSPS) KAKENHI Early-Career Scientists JP18K16292 (to Y.H.), Grant-in-Aid for Scientific Research (B) 20H03668 (to Y.H.), a Research Grant for Young Scholars (to Y.H.), the YASUDA Medical Foundation (to Y.H.), the Uehara Memorial Foundation (to Y.H.), and Medical Research Grants from the Takeda Science Foundation (to Y.H.).

